# Performance at digital testing in Alzheimer’s Disease is predicted by selective disruption of microstructural integrity

**DOI:** 10.64898/2026.02.10.26345988

**Authors:** Sofia Toniolo, Bahaaeddin Attaallah, John Broulidakis, Maria Raquel Maio, Elitsa Slavkova, Shannon Dickson, Olivia Plant, Imran M Idris, Younes Adam Tabi, Christopher Butler, Sian Thompson, Sanjay Manohar, Masud Husain

## Abstract

White matter microstructural abnormalities are increasingly found to be associated with cognitive impairment in Alzheimer’s Disease (AD). Here, we investigated the relationship between visual short-term memory (VSTM) performance, measured using a digital cognitive task, and integrity of brain white matter tracts.

52 AD and 60 age-matched healthy controls were recruited from the Oxford Cognitive Disorders Clinic. An established digital VSTM test – the Oxford Memory Task (OMT) – was used to measure several key memory metrics including: Identification Accuracy (percentage of correctly identified items), Target detection (probability of correctly identifying the target) and Misbinding (erroneously localizing an item to the remembered location of another item in memory). TBSS (Tract-Based Spatial Statistics) was then applied to investigate in which regions of the brain microstructural disruption of physiological diffusivity correlated with behavioural performance.

A key common area, comprising the left optic radiation, forceps major and middle longitudinal fasciculus (MLF), was associated with performance across several VSTM metrics in patients with AD. In addition, misbinding was linked to the left MLF (part III), left inferior fronto-occipital fasciculus and left vertical occipital fasciculus. Microstructural disruption associated with target detection was additionally associated with superior longitudinal fasciculus, and Identification Accuracy with left superior thalamic radiation in AD.

These findings reveal a common shared area of altered diffusivity in AD patients linked to global impairment in VSTM, while distinct types of memory errors are associated with disruption of additional contributions of selective white matter tracts.

**Key points:**

- A key common area, comprising the left optic radiation, forceps major and middle longitudinal fasciculus (MLF), was associated with performance across several visual-short-term memory metrics in patients with Alzheimer’s Disease (AD).
- An analogous area on the right was related to identification accuracy performance in healthy individuals.
- Specific visual working memory functions were associated with selective disruption of other white matter tracts, such as the left MLF (part III), inferior fronto-occipital fasciculus, vertical occipital fasciculus, superior longitudinal fasciculus, and superior thalamic radiation in AD.

## Introduction

Measures of atrophy have long been proven to map biologically to cognition in Alzheimer’s Disease (AD)^1^. However, macroscopic changes occur at late stages during the AD course, when neurodegeneration is already widespread ^2^. As a result, atrophy often becomes apparent late in disease progression. Macroscopic changes are usually preceded by microstructural changes in the brain tissue ^3^, which can be detected before atrophy is overt ^4,5, 6^. Therefore, studying these alterations may be important to fully understand the relationship between underlying pathophysiological processes and cognitive impairment.

Diffusion Tensor Imaging (DTI) is a reliable imaging method used to examine microscopic changes and quantify the breakdown of tissue architecture. Diffusion refers to the random motion of molecules suspended in a medium driven by their thermal energy ^7^. When the molecules are free to move in all directions, this is termed isotropic diffusion, while if they find any anatomical boundary that constrain them to move following a specific direction, the diffusion will be considered anisotropic ^8^. DTI involves fitting of the diffusion property to a tensor model, mathematically represented as an ellipsoid at voxel level. Fractional Anisotropy (FA) is the fraction of the tensor that can be assigned to anisotropic diffusion. It is considered a measure of global WM integrity, with values closer to zero reflecting higher isotropic diffusion, and therefore higher microstructural damage. Axial diffusivity λ1 (axD) represents diffusion along the longer axis of the ellipsoid and is usually considered a proxy of axonal damage. The average of the second and third minor axes (λ2 and λ3) is called radial diffusivity (RD) and reflects diffusivity perpendicular to the major axis of the tensor, likely reflecting myelin damage. Mean diffusivity (MD) is calculated from the average of the three values, indicating the magnitude of overall water diffusion in each voxel, independently of the directionality, indexing overall structural integrity.

Diffusion metrics have been widely studied in AD, for their ability to detect white matter damage even at its earliest stages ^9, 10^. In AD, the physiological neat microstructural milieu is disrupted, leading to a loss of anisotropic diffusion due to degradation of microtubules, loss of axonal structure, demyelination, gliosis and microvascular disarray ^11^. An increase in axD, MD and RD and a decrease in FA have all been widely reported in patients with AD ^12, 13^. Increased Dax and MD are considered to be the first abnormalities to occur, later followed by increased RD and reduced FA ^14, 15, 16^. DTI markers have been found to be altered also in early phases of the disease such as mild cognitive impairment (MCI) and be also able to predict which MCI patients will convert to AD compared to patients whose cognition will remains table over time ^17, 18, 19, 20, 21^. In terms of anatomical localization, while most studies report decreased FA in AD patients especially in the fornix, cingulum and corpus callosum, these alterations are widespread and can be found in several other white matter tracts ^22, 23^.

Although all these different metrics can give us useful information regarding disease progression in AD, brain-behavioral correlations between white matter integrity and cognition are relatively scarce. A recent large-scale study (n = 4467 subjects) has found an association between memory and executive function and diffusion abnormalities, particularly in the fornix and cingulum cortex ^24^. A previous smaller study using the Alzheimer’s Disease Neuroimaging Initiative (ADNI) dataset ^25^ reported that reduced FA correlated with worse cognitive scores on standard tests of cognition, more prominently with memory and executive functions, in a mixed sample of healthy controls and patients with AD ^26^. However, these correlations were not found if the two groups were considered in isolation. One possible explanation could be that these tests are not sensitive enough to detect white matter microstructural deficits in smaller cohorts.

The Oxford Memory Task (OMT) is a visual short-term memory (VSTM) task, which can detect subtle cognitive changes across different neurological conditions, including but not limited to early AD ^27,28,29,30,31,32,33^. Both its tablet version and fully remote online version can discriminate between patients with AD dementia and healthy individuals ^29,30^. It can also aid diagnostic classification in patients with subjective cognitive decline (SCD) and MCI ^31,32^. OMT has been found to outperform standard neuropsychological tests in predicting hippocampal atrophy ^32, 34^. However, data on performance of this task in predicting white matter changes is currently lacking.

VSTM relies on a network of widely distributed brain areas, which suggests a critical role for long-range white matter connections that support information transmission in the brain. Parra et al investigated associations between white matter integrity and VSTM performance in asymptomatic carriers of a causative mutation for familial AD (FAD), symptomatic patients with FAD and healthy controls, deploying a shape-color binding paradigm ^35^. They found that better performance on their VSTM binding task correlated with mean diffusivity values in bilateral genu of the corpus callosum (corresponding to the Forceps Minor), left Superior Longitudinal Fasciculus (SLF), and left hippocampal region of the Cingulum. FA values did not significantly predict task performance in that study, and these correlations were found only in symptomatic FAD patients. However, that study deployed an a priori region of interest (ROI) approach, based on ROIs primarily involved in FAD, and did not explore brain-behavioural correlations in the whole brain.

In this study we conducted a whole brain analysis to investigate which areas of the brain were associated with Oxford Memory Task metrics. Tract-based spatial statistics (TBSS) is an ideal method to answer this question as, similarly to Voxel Based Morphometry (VBM) for grey matter, allows the voxelwise statistics to be calculated between each voxel and behavioural measure directly. Crucially, TBSS is better than VBM when using diffusion imaging data, as overcomes problems in alignment between different subjects’ diffusion images and in spatial smoothing (local averaging of noise used in cross-subject registration)^36^. Moreover, it is a robust and validated tool in AD patients ^37, 38^. We investigated whether successful retrieval, binding abilities and reaction times, associated with different white matter tracts in a cohort of elderly healthy controls (EHC) and AD patients.

## Methods

### Participants

52 AD and 60 EHC were recruited from the Cognitive Disorders Clinic at the John Radcliffe Hospital in Oxford and through open day events respectively. EHC were > 50 years old, had no reported psychiatric or neurological illness, were not on psychoactive drugs and scored above the cut-off for normality at standard tests of cognition (88/100 total at the Addenbrooke’s Cognitive Examination (ACE-III) ^39^. AD patients were defined as per Jack et al 2018 ^65^ as having Alzheimer’s clinical syndrome and hereinafter referred to as the AD group. They were diagnosed according to their clinical phenotype, supporting imaging (magnetic resonance imaging (MRI) and fluorodeoxyglucose positron emission tomography (FDG-PET)) and Cerebrospinal fluid (CSF)/plasma biomarkers if available. AD patients were enrolled in the study if their total ACE-III score was higher than 45/100. Participants underwent the OMT task, a standard neuropsychological test, i.e., the ACE-III, and a 3T MRI brain scan. MRI scans were visually inspected independently by 2 trained neurologists (S.T. and I.I.), and subjects with significant white matter burden (Fazekas Score ^40^ = or > 1) were excluded from the study.

The study was performed in accordance with the ethical standards as laid down in the 1964 Declaration of Helsinki and its later amendments. Ethical approval was granted by the University of Oxford ethics committee (IRAS ID: 248379, Ethics Approval Reference: 18/SC/0448). All participants gave written informed consent prior to the start of the study.

### Oxford Memory Task

The task design of OMT has been extensively previously described elsewhere ^28,29,30,31,32,33^. In brief, participants were presented with either 1 or 3 fractals located randomly on the screen. They were asked to remember the identity of the fractals and their locations. After a delay of either 1 or 4 seconds, two fractals appeared at the centre of the screen along the vertical axis. One of these had appeared in the memory array (target) whereas the other one was a foil, which had not been shown in the current trial. Participants were required to touch the target and drag it to its original location. The Mixture Model of working memory by Bays et al ^14^ was then fitted to the data to unravel the differential contribution of memory errors to our dataset. Different working memory metrics extracted included:

- **Identification Time**: the time in seconds taken to identify the correct object.
- **Localization Time**: the time in seconds to drag the chosen object to its remembered location.
- **Identification Accuracy**: the proportion of trials in which participants correctly identified the target.
- **Target detection:** the probability of correctly identifying the target.
- **Misbinding**: the probability of mislocalizing a correctly identified item to the remembered location of another item in the memory array.
- **Guessing**: the probability of random guessing responses.

### Statistical analysis (demographics and neuropsychological tests)

Statistical significance was derived from either independent sample t-test or Mann*–*Whitney U test according to the normality of the data. χ^2^ test was used to compare gender distribution across groups. Rank-Biserial correlation was used as a measure of effect size for Mann*–*Whitney U derived statistics.

### MRI protocol

MRI was performed at 3T (Siemens Magnetom, Verio syngo MR B17) with the following sequences, harmonized to the UK Biobank protocol ^50, 51^: T1-weighted sequence (3D MPRAGE, sagittal, TR = 2000 ms, TE = 1.94 ms, TI = 880 ms, in-plane acceleration factor (R) = 2, flip angle 8°, voxel-size = 1.0 × 1.0 × 1.0 mm, 208 × 256 × 256 matrix), T2-FLAIR sequence (FLAIR, 3D SPACE, sagittal, TR = 5000 ms, TE = 397 ms, TI = 1800 ms, partial fourier (PF) = 7/8, fatsat, voxel-size = 1.05 × 1.0 × 1.0 mm, 192 × 256 × 256 matrix), Diffusion-weighted images (dMRI) (Multiband acceleration factor (MB) = 3, TR = 3600 ms, TE = 92 ms, PF = 6/8, fatsat, voxel size 2.0 × 2.0 × 2.0 mm, 104 × 104 × 72 matrix) with opposite phase encoding directions (AP and PA) to allow for more robust distortion correction (AP: 104 directions, diffusion weighting: b1=0 s/mm^2^, b2=2000 s/mm^2^ and PA: 7 directions, diffusion weighting: b1 = 0 s/mm^2^, b2 = 1000 s/mm^2^), Resting-state fMRI (rsfMRI) images (MB = 8, flip angle 52°, fat sat, TR = 735 ms, TE = 39 ms, voxel size = 2.4 × 2.4 × 2.4 mm, 88 × 88 × 64 matrix).

### MRI pre-processing

Firstly, nonlinear registration from all FA images to a target image was performed, followed by an affine alignment to a standard space, the 1×1×1 mm Montreal Neurosciences Institute (MNI152) space, to create a mean FA mask. Secondly, the mean FA mask was skeletonized, where the skeleton represents the centre of all tracts (as delineated on an FA map), which are common to all subjects. The mean FA skeleton was subsequently thresholded to restrict the analysis to voxels that have been successfully aligned across subjects, creating a binarized skeleton mask. Each subject’s FA was projected to the main skeleton, and a 4D image containing the projected skeletonized FA data was produced.

### MRI statistical analysis

Voxelwise analysis was carried out within the skeletonized FA data, using the *randomise* function of FSL. A group contrast between EHC and AD was first performed, examining areas in which AD had higher (AD > EHC) or lower (AD < EHC) FA compared to healthy controls, to check the extent of diffusivity damage in our sample. Subsequently, we tested brain-behavioural correlations by applying a voxelwise general linear model (GLM), using permutation-based non-parametric testing (n permutations = 5000) for correcting for multiple comparisons across space. Correlations between the eight VSTM metrics and FA were calculated in the whole sample (EHC and AD), and separately for each group (EHC, AD), and whether the VSTM metric was associated positively or negatively FA. Age, gender and education were included in the GLM as covariates. All correlation analyses between VSTM metrics and FA were restricted within the group difference mask derived from the first contrast (AD < HC). Results reported have as minimum cluster size of n = 10, considered to provide a good balance of Types I and II error rates ^41^. The XTRACT HCP Probabilistic Tract Atlas was used as reference labelling system for white matter tracts ^42^. Alternative notations from the other JHU white matter atlases^43^ have also been included, particularly if no match was found within a specific tract and the XTRACT atlas. Finally, for each correlation the probability of the overall cluster of belonging to the tracts within the XTRACT atlas was extracted using FSL *atlasquery* function.

## Results

### Visual short term memory performance

Patients’ basic demographics and tests results are presented in **Table 1**. The two groups did not differ significantly in terms of age, but there was a higher proportion of males and lower education levels in patients with AD compared to EHC. As expected, performance on the ACE-III and the OMT was lower in AD compared to EHC, across all the eight VSTM metrics.

**Table 1.** Demographics and tests. M = male, F = female. Addenbrooke’s Cognitive Examination (ACE-III).

|  | **EHC**<br>(n = 60) | **AD**<br>(n = 52) | **p-value** | **Rank-Biserial**<br>**Correlation** |
| --- | --- | --- | --- | --- |
| **Age** | 66.45 (7.9) | 68.3 (10.1) | 0.465 |  |
| **Gender (M/F)** | 22/38 | 30/22 | *0.026 |  |
| **Education** | 17.3 (6.5) | 14.6 (4.2) | *0.005 |  |
| **ACE-III** | 97.3 (2.5) | 74.6 (18.1) | *<0.001 |  |
| **Identification time** | 1.89 (0.4) | 3.14 (1.6) | *<0.001 | -0.590 |
| **Localization time** | 3.8 (0.8) | 6.3 (3.6) | *<0.001 | -0.517 |
| **Identification accuracy** | 0.93 (0.0) | 0.81 (0.1) | *<0.001 | 0.765 |
| **Target detection** | 0.76 (0.1) | 0.56 (0.1) | *<0.001 | 0.793 |
| **Guessing** | 0.13 (0.05) | 0.28 (0.1) | *<0.001 | -0.789 |
| **Misbinding** | 0.21 (0.7) | 0.32 (0.1) | *<0.001 | -0.703 |

### Voxel-wise correlations between VSTM metrics and white matter tracts across the entire sample (EHC and AD)

Higher Misbinding, higher Guessing and lower Target detection were associated with reduced FA in the same key region, which encompassed three main tracts: the left Optic Radiation (OR), the left Forceps Major (FM) and the left Middle Longitudinal Fasciculus (MLF) (**Figure 1**, **Table 2** and **3**). Additionally, for all three of these metrics there was involvement, to a much lesser extent, of the left Inferior Fronto-occipital Fasciculus (Inf-FOF) and the left Vertical Occipital Fasciculus (VOF). Misbinding was associated with the smallest cluster size and had higher involvement of the Optic Radiation compared to Guessing and Target detection (**Figure 1** and **Table 3**), reflected also by the peak being within the left Optic Radiation (**Table 2**).

**Figure 1.**
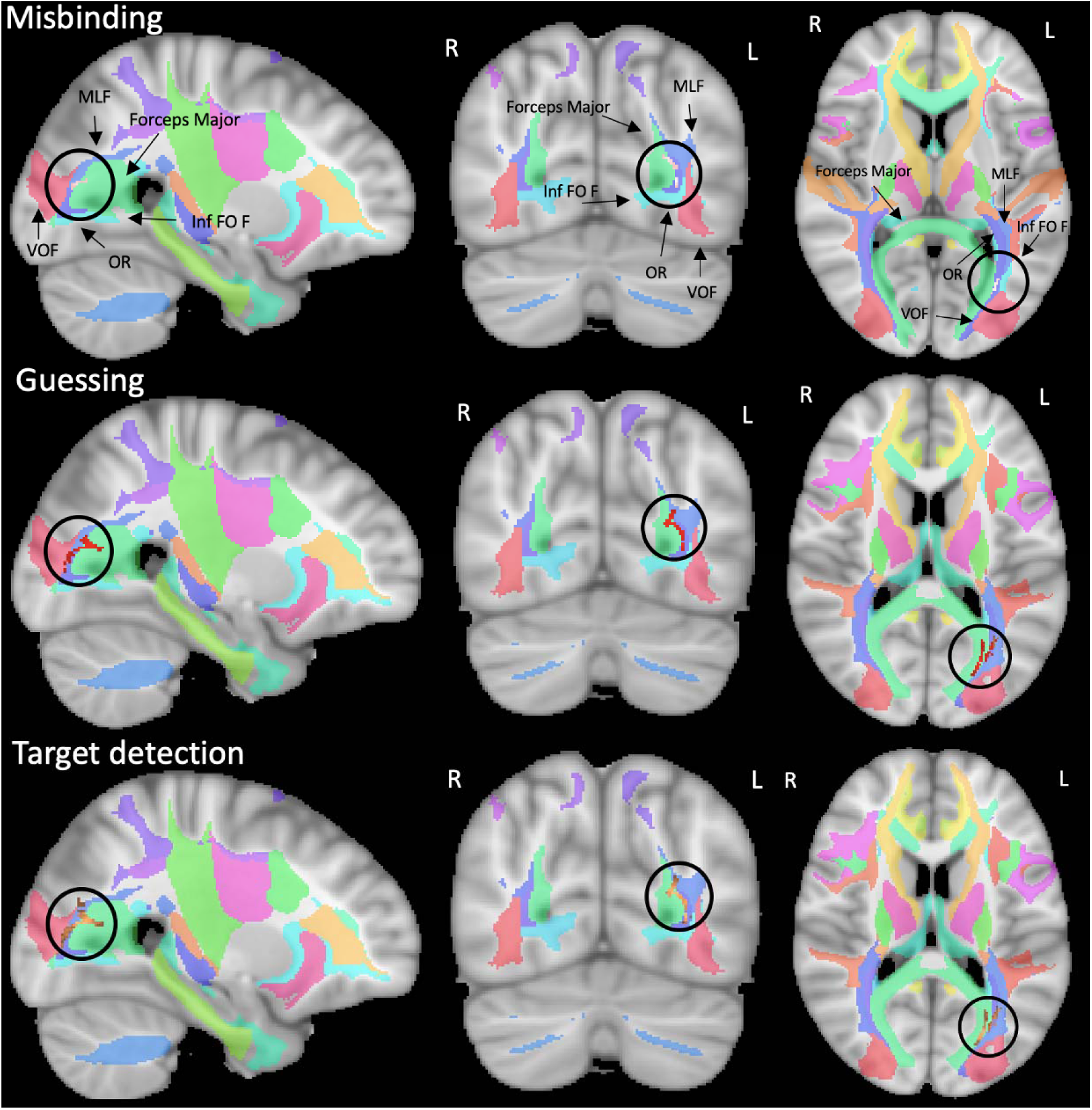
Correlations between VSTM metrics (mixture model metrics) and FA across subjects. Misbinding in pink (upper row), Guessing in red (middle), Target Detection in copper (bottom). From the XTRACT atlas: Optic Radiation (OR) in violet, Forceps major in mint green, Middle Longitudinal Fasciculus (MLF) in light violet, Inferior Fronto-occipital Fasciculus (inf FOF) in light blue, Vertical Occipital Fasciculus (VOF) in coral red. R = right hemisphere, L = left hemisphere.

**Table 2.** Correlations between VSTM metrics and FA in AD and EHC: Clusters and diffusivity peaks. L = left. JHU = Johns Hopkins University atlas.

|  | N<br>cluster<br>s | Xtract atlas | Cluster<br>Size | Coordinates<br>(x,y,z) |  |  | Z-score |
| --- | --- | --- | --- | --- | --- | --- | --- |
| **Misbinding** | 1 | Optic radiation (L) | 134 | -31 | -70 | 6 | 4.1 |
| **Guessing** | 1 | Forceps major | 423 | -24 | -74 | 14 | 3.77 |
| **Target detection** | 1 | Forceps major | 614 | -26 | -73 | 13 | 3.79 |
| **Identification time** | 1 | Vertical occipital Fasciculus (L) | 806 | -34 | -78 | 16 | 4.65 |
|  | 2 | Superior Longitudinal Fasciculus 1 (L) | 276 | -15 | -58 | 53 | 3.38 |
|  | 3 | Superior Longitudinal Fasciculus 1 (L) | 66 | -22 | -66 | 32 | 3.46 |
|  | 4 | Superior Thalamic Radiation (L) | 42 | -18 | -12 | 39 | 38.7 |
| **Identification accuracy** | 1 | Superior Longitudinal Fasciculus 2 (L) | 1635 | -33 | -59 | 26 | 3.55 |
|  | 2 | Cingulum - posterior division (L) (JHU) | 1083 | -16 | -60 | 35 | 3.72 |
|  | 3 | Corticospinal Tract (L) | 268 | -23 | -28 | 42 | 3.82 |

**Table 3.** Correlations between VSTM metrics and FA in AD and EHC: Tracts probability. MB = Misbinding, GS = Guessing, TG = Target detection, IT = Identification time, IA = Identification accuracy. L = left.

| White matter tract | MB | GS | TG | IT | IA |
| --- | --- | --- | --- | --- | --- |
| Optic Radiation (L) | 47.6884 | 33.2501 | 37.4603 | 10.9997 | 10.8108 |
| Forceps major | 28.7136 | 37.2356 | 30.8491 | 16.9285 | 7.9668 |
| Middle Longitudinal Fasciculus (L) | 20.7313 | 23.0339 | 25.0432 | 9.7308 | 6.7542 |
| Inferior Fronto-occipital Fasciculus (L) | 7.626 | 4.2938 | 6.3895 | 0.977 | 3.7629 |
| Vertical Occipital Fasciculus (L) | 1.9992 | 5.0412 | 7.5466 | 4.3322 | 2.0733 |
| Superior Longitudinal Fasciculus 1 (L) |  |  |  | 4.4798 | 4.5177 |
| Superior Longitudinal Fasciculus 2 (L) |  |  |  | 4.0323 | 3.025 |
| Superior Longitudinal Fasciculus 3 (L) |  |  |  | 0.8422 | 1.1784 |
| Superior Thalamic Radiation (L) |  |  |  | 0.7481 | 0.8037 |
| Corticospinal Tract (L) |  |  |  |  | 1.1649 |
| Arcuate Fasciculus (L) |  |  |  |  | 0.7639 |

Higher Identification time and Identification accuracy were correlated with lower FA in the same core area as the three mixture model metrics, and in additional tracts including the left Superior Longitudinal Fasciculus (SLF) (1,2,3) and left Superior Thalamic Radiation (STR) (**Figure 2**, **Table 2** and **3**). Higher Identification accuracy was associated with reduced FA also in the left Corticospinal Tract (CST), left Arcuate Fasciculus (AF) and the left Cingulum (**Figure 2**, **Table 2** and **3**).

**Figure 2.**
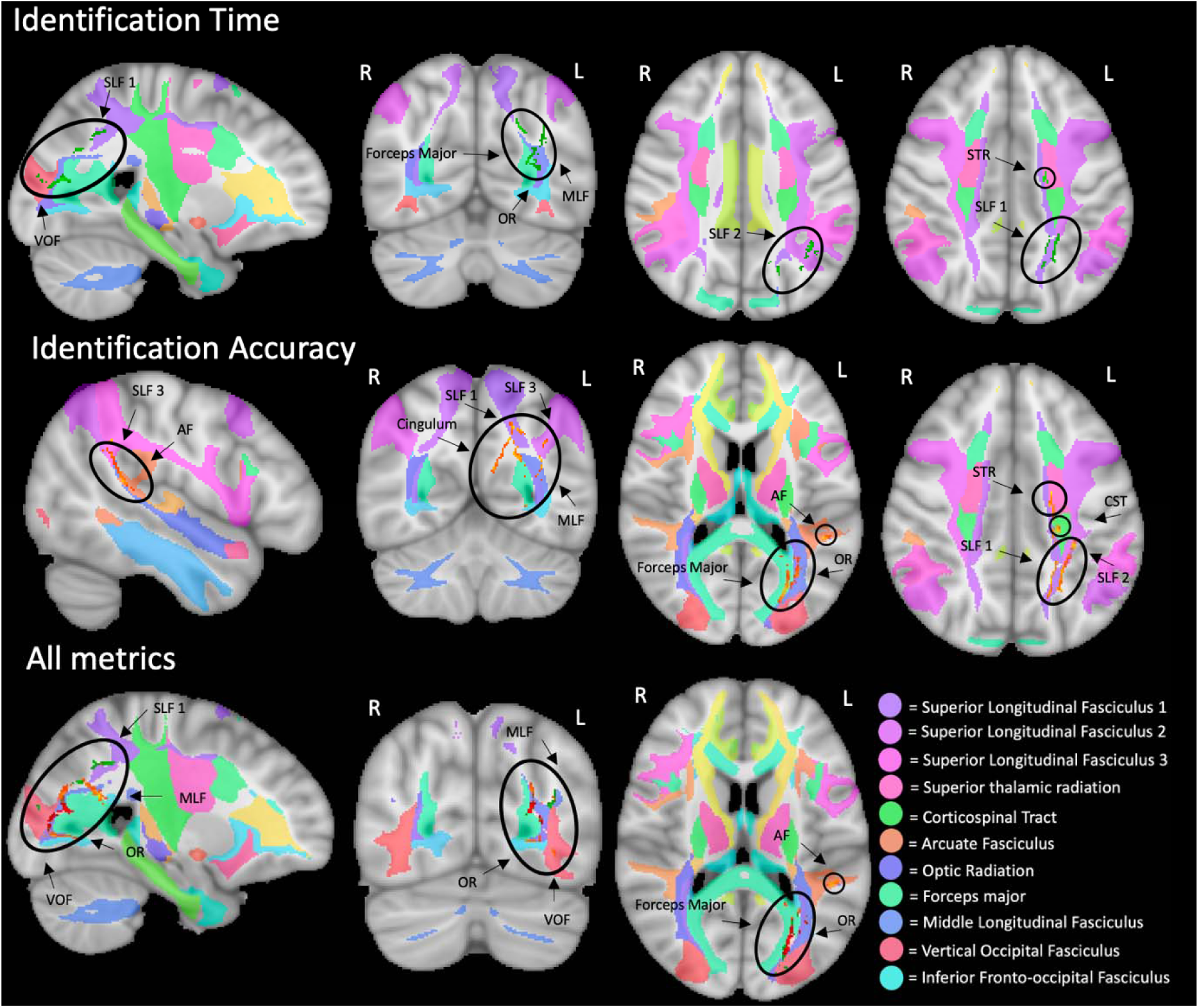
Identification metrics and correlations between all VSTM metrics and FA. Identification Time in emerald green (upper row), Identification Accuracy in orange (middle row), significant clusters from all metrics are overlaid in the bottom row, with the same color coding as Figure 7. From the XTRACT atlas: Superior Longitudinal Fasciculus 1 (SLF) in lilac, SLF 2 in purple, SLF 3 in light purple, Superior Thalamic Radiation (STR) in pink, Corticospinal Tract (CST) in acid green, Arcuate Fasciculus (AF) in orange, Optic Radiation (OR) in violet, Forceps major in mint green, Middle Longitudinal Fasciculus (MLF) in light violet, Inferior Fronto-occipital Fasciculus (Inf-FOF) in light blue, Vertical Occipital Fasciculus (VOF) in coral red.

All metrics displayed the expected effect direction, with lower Target detection and Identification accuracy and higher Misbinding, Guessing and Identification Time, being associated with reduced FA. No areas were found in the reverse contrast for any metric.

### Correlations between VSTM metrics and white matter tracts in each cohort (AD and EHC)

Next, we investigated whether these results held true in the cohort of AD patients alone. In this group, higher Misbinding and higher Guessing were associated with reduced FA in the same key regions observed in the previous analysis, including the left Optic Radiation, the Forceps Major and the left Middle Longitudinal Fasciculus and to a lesser extent, the left Inferior Fronto-occipital Fasciculus and the left Vertical Occipital Fasciculus (**Figure 3**, **Table 4** and **5**), with the difference that in this case Guessing had the smallest cluster size. However, this analysis confirmed that, similarly to the previous analysis, Misbinding had a much higher involvement of the left Optic radiation compared to Guessing, while higher Guessing was associated more tightly with lower FA in the Forceps major (**Figure 3**). This was supported also by the localization of the respective peaks and from the distribution of tracts’ probability (**Table 4** and **5**). Unlike the analysis in the whole group, in AD patients alone Target detection not only involved these core areas but also showed a significant involvement of the left Superior Longitudinal Fasciculus 1 and 2, posterior Cingulum and of a small cluster in the right Inferior fronto-occipital Fasciculus (**Figure 3**, **Table 4** and **5**).

**Figure 3.**
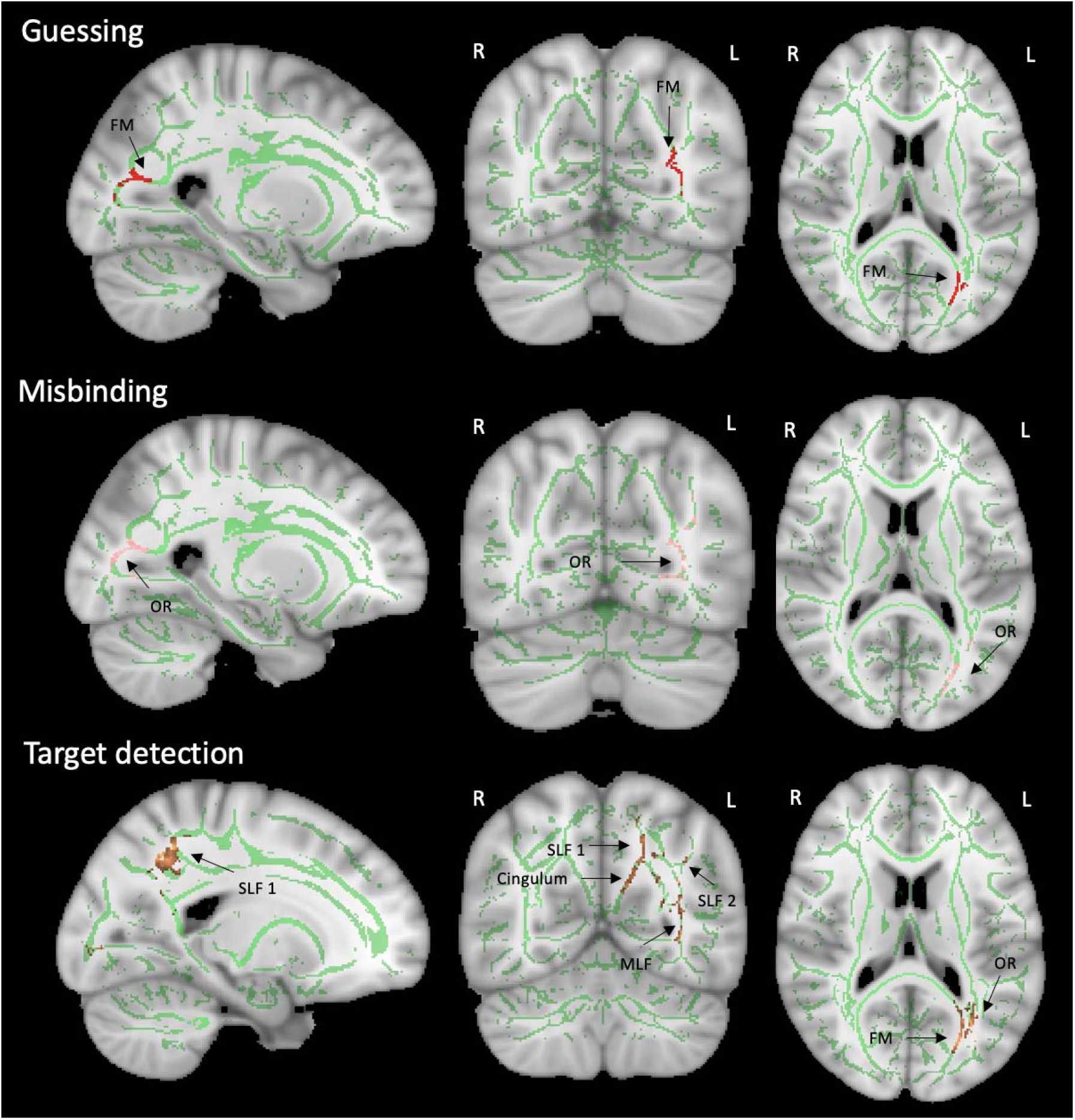
Correlations between mixture model VSTM metrics and FA in AD. Guessing in red (upper row), Misbinding in pink (middle), Target Detection in copper (bottom). From the XTRACT atlas: Optic Radiation (OR), Forceps major (FM), Middle Longitudinal Fasciculus (MLF), Superior Longitudinal Fasciculus (SLF). R = right hemisphere, L = left hemisphere.

**Table 4.**
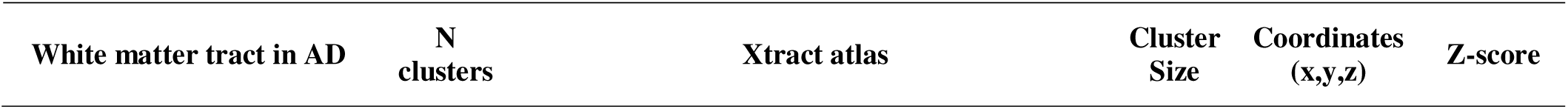
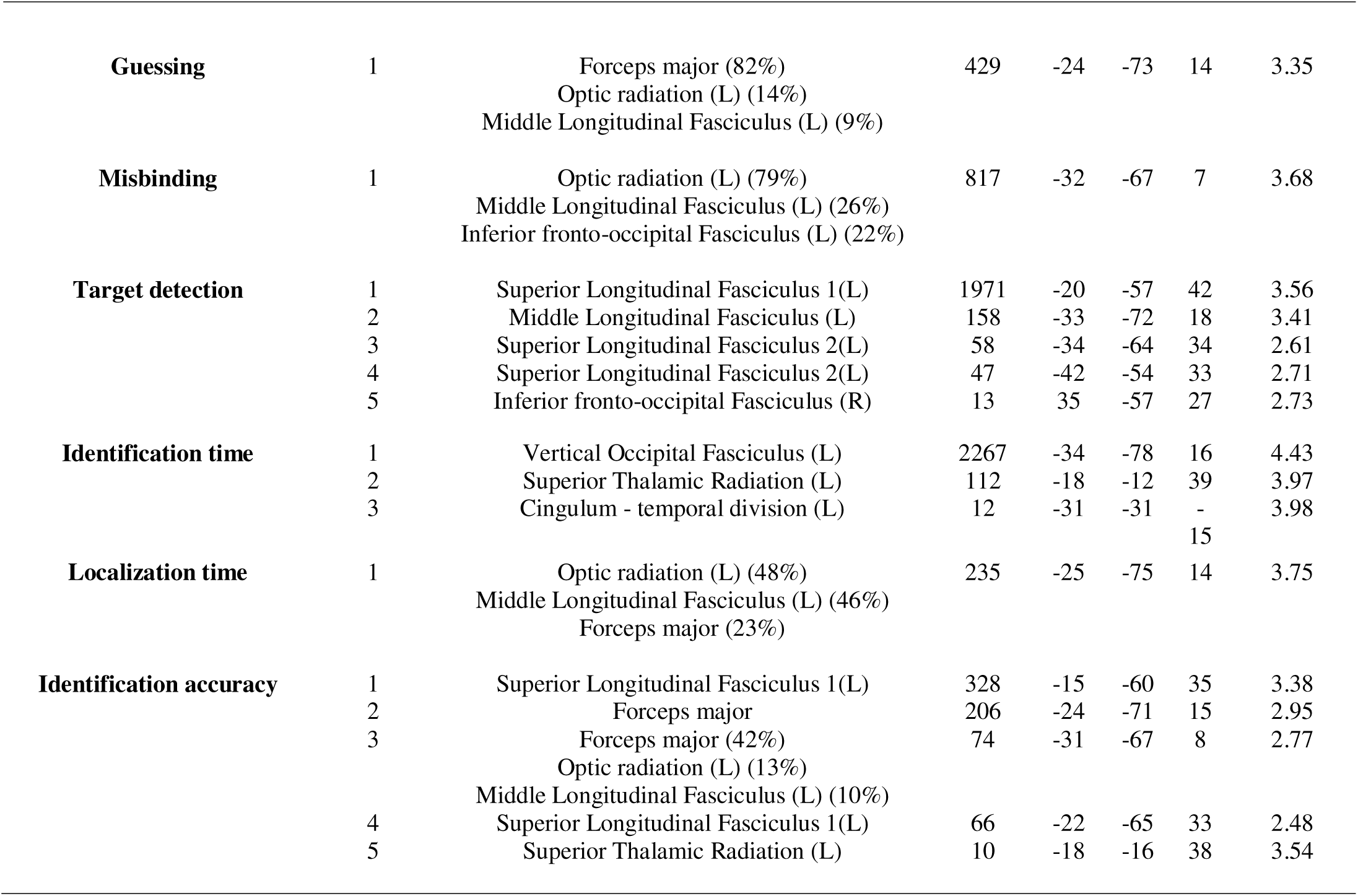
Correlations between VSTM metrics and FA in AD: Clusters and diffusivity peaks. L = left, R = right.

In the AD cohort, Identification time had the biggest cluster across all VSTM metrics, with longer times correlating with lower FA in the same areas as Target detection, but the peak of the main cluster this time being in the left Vertical Occipital Fasciculus (**Figure 4**, **Table 4** and **5**). Additionally, reduced FA was correlated with higher Identification time in the left Superior Thalamic Radiation and in a small cluster in the left temporal section of the Cingulum (**Figure 4**, **Table 4** and **5**). Unlike the previous analysis in the whole cohort, in AD patients there was also a small area that was associated with longer Localization times, corresponding to the key area found in Guessing and Misbinding, with a prominent involvement of the Forceps Major (**Figure 4**, **Table 4** and **5**).

**Figure 4.**
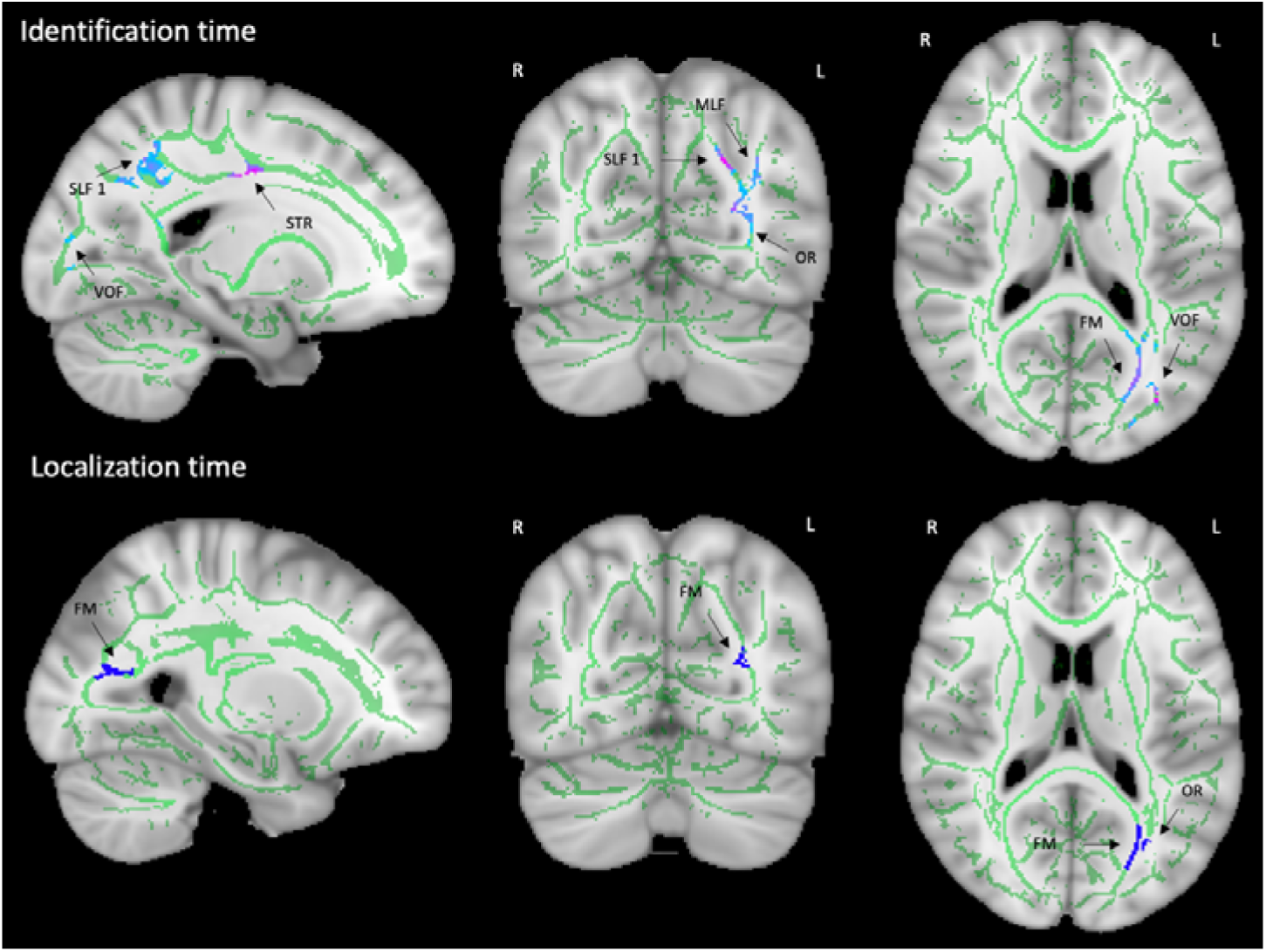
Correlations between reaction times metrics (Identification time and Localization time) and FA in AD. Identification time in cool colours, the pinker the higher the Z-score (upper row), Localization time in blue (lower row). Superior Longitudinal Fasciculus (SLF), Superior Thalamic Radiation (STR), Vertical Occipital Fasciculus (VOF), Middle Longitudinal Fasciculus (MLF), Forceps major (FM), Optic Radiation (OR). R = right hemisphere, L = left hemisphere.

**Table 5.**
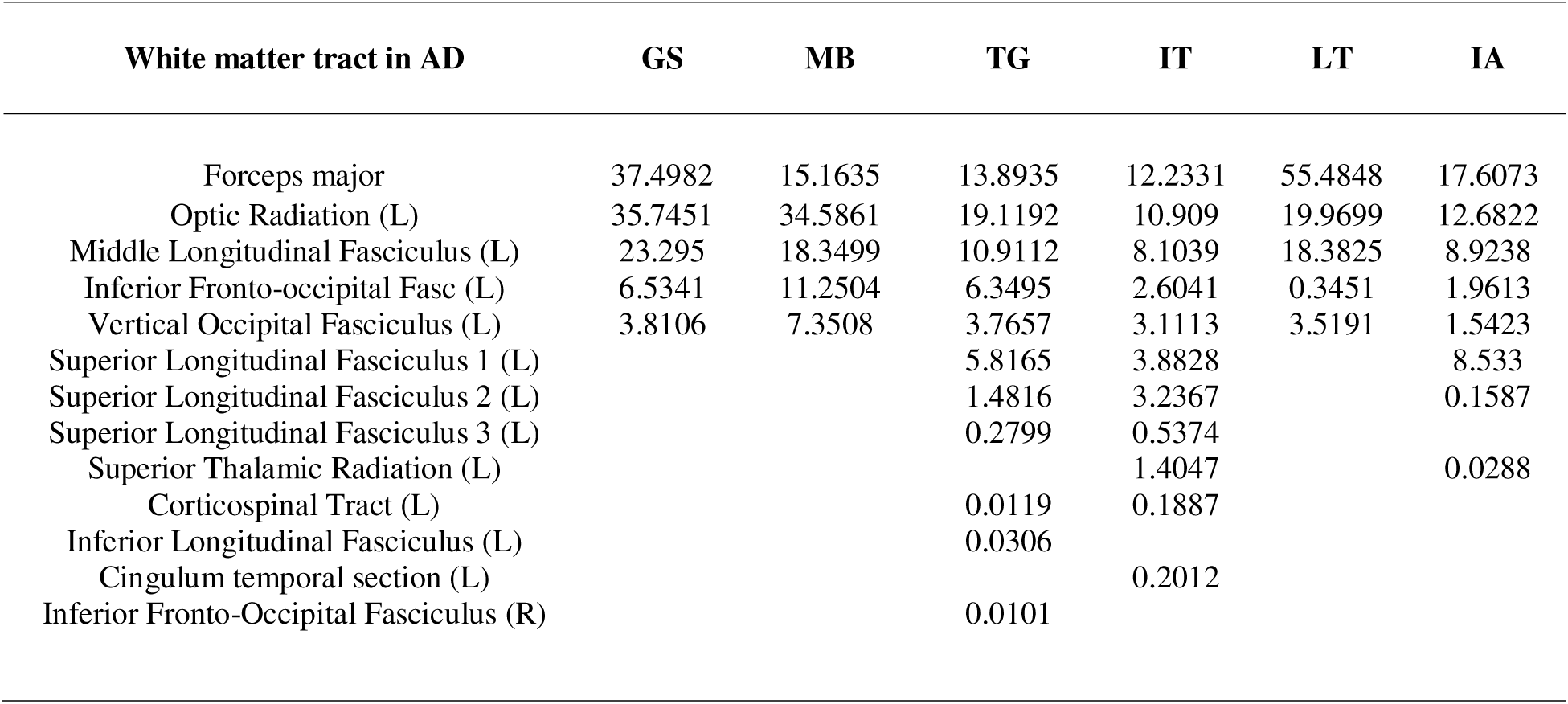
Correlations between VSTM metrics and FA in AD: Tracts probability. MB = Misbinding, GS = Guessing, TG = Target detection, IT = Identification time, LT = Localization time, IA = Identification accuracy. L = left.

| White matter tract in AD | GS | MB | TG | IT | LT | IA |
| --- | --- | --- | --- | --- | --- | --- |
| Forceps major | 37.4982 | 15.1635 | 13.8935 | 12.2331 | 55.4848 | 17.6073 |
| Optic Radiation (L) | 35.7451 | 34.5861 | 19.1192 | 10.909 | 19.9699 | 12.6822 |
| Middle Longitudinal Fasciculus (L) | 23.295 | 18.3499 | 10.9112 | 8.1039 | 18.3825 | 8.9238 |
| Inferior Fronto-occipital Fasc (L) | 6.5341 | 11.2504 | 6.3495 | 2.6041 | 0.3451 | 1.9613 |
| Vertical Occipital Fasciculus (L) | 3.8106 | 7.3508 | 3.7657 | 3.1113 | 3.5191 | 1.5423 |
| Superior Longitudinal Fasciculus 1 (L) |  |  | 5.8165 | 3.8828 |  | 8.533 |
| Superior Longitudinal Fasciculus 2 (L) |  |  | 1.4816 | 3.2367 |  | 0.1587 |
| Superior Longitudinal Fasciculus 3 (L) |  |  | 0.2799 | 0.5374 |  |  |
| Superior Thalamic Radiation (L) |  |  |  | 1.4047 |  | 0.0288 |
| Corticospinal Tract (L) |  |  | 0.0119 | 0.1887 |  |  |
| Inferior Longitudinal Fasciculus (L) |  |  | 0.0306 |  |  |  |
| Cingulum temporal section (L) |  |  |  | 0.2012 |  |  |
| Inferior Fronto-Occipital Fasciculus (R) |  |  | 0.0101 |  |  |  |

Lastly, in the AD cohort, lower Identification accuracy was associated with lower FA in the same key areas involved in Misbinding and Guessing, and additionally the left Superior Longitudinal Fasciculus 1 and the left Superior Thalamic Radiation (**Figure 5**, **Table 4** and **5**).

**Figure 5.**
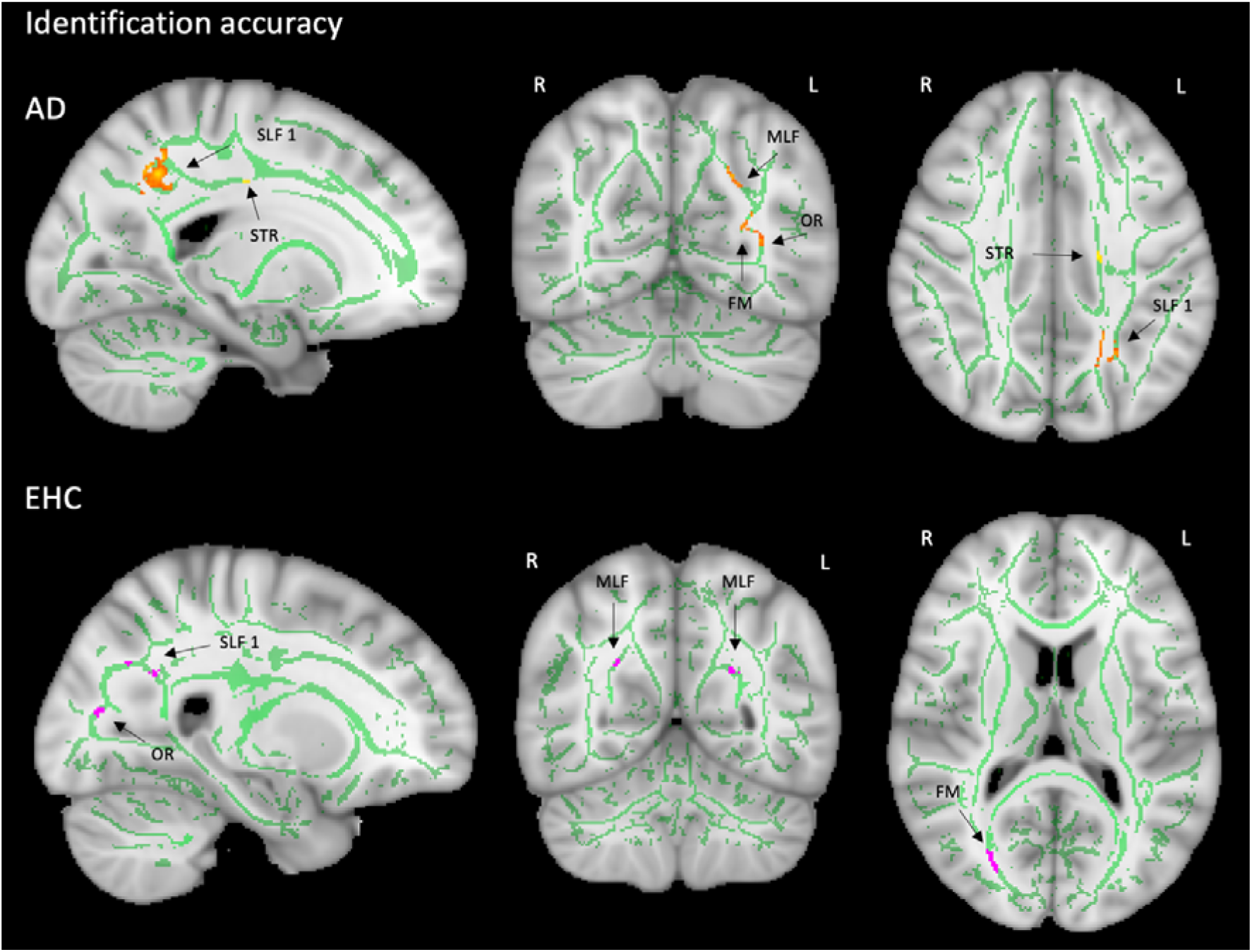
Correlations between Identification accuracy and FA in AD and EHC. Identification accuracy in orange for AD (top row), and in magenta for EHC (lower row). Superior Longitudinal Fasciculus (SLF), Superior Thalamic Radiation (STR), Middle Longitudinal Fasciculus (MLF), Forceps major (FM), Optic Radiation (OR). R = right hemisphere, L = left hemisphere.

All metrics displayed the expected effect direction, as in the previous analysis, with lower Target detection and Identification accuracy and higher Misbinding, Guessing, Identification and Localization Time, being associated with reduced FA. No areas were found in the reverse contrast for any metric in AD patients.

We did not find any association between VSTM metrics and white matter diffusivity changes in EHC, except for Identification accuracy. Notably, in EHC, lower Identification accuracy was correlated with lower FA in the right hemisphere, in a homologous region compared to the key common area across VSTM metrics seen on the left in AD patients (including Forceps major, Optic Radiation and Middle Longitudinal Fasciculus), (**Figure 5**, **Table 6** and **7**). Additionally, there was an involvement of the right Superior Longitudinal Fasciculus 1, and the left Middle Longitudinal Fasciculus.

**Table 6.** Correlations between VSTM metrics and FA in EHC: Clusters and diffusivity peaks.

| White matter tract in EHC | N clusters | Xtract atlas | Cluster Size | Coordinates (x,y,z) |  |  | Z-score |
| --- | --- | --- | --- | --- | --- | --- | --- |
| Identification accuracy | 1 | Forceps major (83%)<br>Optic radiation (R) (43%)<br>Middle Longitudinal Fasciculus (R) (24%) | 200 | 32 | -67 | 4 | 3.49 |
|  | 2 | Superior Longitudinal Fasciculus 1 (R) | 42 | 24 | -74 | 36 | 4.77 |
|  | 3 | Middle Longitudinal Fasciculus (R) | 26 | 25 | -56 | 28 | 3.36 |
|  | 4 | Middle Longitudinal Fasciculus (L) | 24 | -25 | -56 | 24 | 4.2 |

**Table 7.** Correlations between VSTM metrics and FA in EHC: Tracts probability. IA = Identification accuracy

| White matter tract in EHC | IA |
| --- | --- |
| Optic Radiation (R) | 34.4549 |
| Forceps major | 13.4101 |
| Middle Longitudinal Fasciculus (R) | 16.5652 |
| Inferior Fronto-occipital Fasciculus (R) | 6.5458 |
| Vertical Occipital Fasciculus (R) | 10.9140 |
| Superior Longitudinal Fasciculus 1 (R) | 0.1026 |
| Middle Longitudinal Fasciculus (L) | 0.2234 |
| Optic Radiation (L) | 0.0241 |

## Discussion

In this study whole brain TBSS analysis was deployed to investigate voxelwise associations between white matter integrity and the VSTM metrics derived from the OMT, in a cohort of sporadic late onset AD patients and elderly healthy controls. Across the entire cohort, and particularly in patients with AD, the analysis revealed a key white matter area in the left hemisphere to be associated with several VSTM metrics: Guessing, Misbinding, Target Detection, Identification Time, Localization Time and Identification accuracy. It consisted of left OR, Forceps major, MLF, and to a much lesser extent the IFOF and VOF (**Figure 1-5**).

Significant brain-behavioural correlations between FA in several white matter tracts and Identification accuracy on the VSTM task, primarily in the right hemisphere, were found in healthy elderly adults (**Figure 5**). These tracts included on the right the Forceps Major, OR, SLF-1 and bilaterally the MLF. The results are encouraging as even by using an easily accessible measure such as FA, it was possible to detect a preferential involvement of specific white matter tracts in VSTM abilities in elderly healthy controls.

The OR (known also as posterior thalamic radiation) is a white matter bundle connecting the primary visual cortex to the lateral geniculate nucleus ^44^. Integrity of the right OR has previously been associated with performance on a VSTM task (Visual Pattern Test, Juelich version) ^45^ in elderly adults, but this is the first time that we were able to validate this association across multiple VSTM metrics, deploying the mixture model of working memory. The left OR is severely affected in AD, even in the earliest stages of the disease ^46^. Indeed, reduced FA in the left OR is considered to be amongst the best predictors for early discrimination between healthy controls, patients with subjective and mild cognitive impairment, but also for clinical progression to AD dementia ^18, 46^. In AD it has also been associated with worse performance on multiple standard neuropsychological tests including the mini mental state examination (MMSE), clinical dementia rating scale sum of boxes (CDR-sob), the Alzheimer’s Disease Assessment Scale - Cognitive Subscale (ADAS-cog) and Rey Auditory Verbal Learning Test ^18, 46^. The left OR has also been linked to the precision of color representation in a VSTM task in patients with stroke ^47^. However, it is the first time that the disruption of microstructural integrity in the left OR in AD has been associated with worse performance across multiple VSTM metrics.

Similarly, the FM, a white matter tract connecting the occipital lobes bilaterally, is heavily affected in AD across all disease stages, and shows a very tight correlation with measures of cognitive impairment ^18, 46^. DTI changes in the FM have been found to be the earliest markers of microstructural disruption in patients with FAD, becoming apparent a decade before symptom onset ^48^.

Our results suggest that the integrity of the OR and the FM is an important predictor of VSTM performance, but this appears to vary across physiological and pathological ageing, with the right hemisphere being more important in healthy elderly controls, as opposed to the left hemisphere in AD patients. Moreover, whilst these two structures were associated with multiple VSTM metrics, Misbinding might have a more prominent involvement of the left OR, whilst higher Guessing seems more tightly associated with disruption of the FM, which is shown by the peaks and the percentage of tracts involvement across the whole cohort and particularly in AD patients (**Figure 1-3**, **Table 2-5**). We postulate that an increasingly faulty communication between the two occipital lobes (carried by the FM), causes spatial information of the object-to-be-remembered to be compromised. Conversely, misbinding errors might arise from a disrupted connection from the occipital lobe, through the medial temporal lobe to the thalamus, where some spatial information might be retained but wrongly processed as linked to another object in memory.

The MLF is far less studied, as its existence in humans has been unravelled relatively recently by the use of DTI ^49^. It connects the angular gyrus to the superior temporal gyrus, and whilst its functions have not been completely understood, it has been proposed as being likely associated with a traditional right – left lateralization for visuospatial processing in the right hemisphere, and speech in the left hemisphere ^49, 50^. Our data might shed a light on the differential function of the MLF in health and disease. In healthy subjects the integrity of both the right and the left MLF is important for correctly identifying an item, measured by Identification accuracy of our VSTM task (**Figure 5**, **Table 6 and 7**). On the other hand, in AD patients only the left MLF seems to be important to guarantee optimal performance across different VSTM metrics.

Two white matter tracts that have been minimally but homogeneously involved in VSTM performance at our task were the IFOF and the VOF. The IFOF is a tract that connects directly occipital and frontal lobes, it originates from the lingual, posterior fusiform, cuneus and polar occipital cortex, passes through the internal capsule, and terminates in the inferior frontal gyrus, medial fronto-orbital region and frontal pole ^51^. The IFOF has been associated with visual working memory performance in healthy individuals, and particularly with visual learning, and is thought to be negatively impacted by ageing ^52^. Performance on a VSTM task in a cohort of healthy controls and patients with multiple sclerosis, showed that the Inf-FOF was a strong predictor of performance for object but not for spatial localization accuracy in both groups ^53^. Our data support the idea that it is not associated with a specific VSTM function, but the integrity of the left IFOF supports general VSTM functioning in patients with AD.

The VOF is the only white matter tract connecting dorsolateral and ventrolateral visual cortex ^54^. It is thought to be crucial to integrate ventral regions, that are important for the perception of visual stimuli, and dorsal regions, involved in the control of eye movements, attention, and motion perception ^54^. Our data support that its disruption contributes to overall VSTM deficits in AD patients and particularly seems to have the highest association (highest peak) with Identification time (**Figure 4**, **Table 4**). This might reflect the complex interaction between visual perception and the execution of the movement needed to select the correct object, which can be degraded in patients with AD, leading to longer Identification times.

The left SLF has been shown to be associated with Identification accuracy and Identification time across the whole cohort and additionally with Target detection in patients with AD (**Figure 1-5**, **Table 2-5**), while the right SLF 1 was associated with Identification accuracy in our healthy elderly controls (**Figure 5**, **Table 6 and 7**). The SLF can be divided into dorsal (SLF 1), middle (SLF 2), and ventral sections (SLF 3), with different patterns connections to other brain regions and functional specialization ^55, 56^. According to these studies, the SLF 1 connects the superior parietal lobule to superior frontal lobe (motor and premotor areas) and has been associated with higher order motor control. Conversely, the SLF 2 conveys information from the inferior parietal lobe (particularly, the intraparietal sulcus) to the prefrontal cortex (as the superior and middle frontal gyrus) and is particularly involved in visual space perception and re-orienting of spatial attention.

Finally, the SLF 3 connects the rostral part of the inferior parietal lobule with the inferior frontal gyrus and has been associated with monitoring orofacial and hand actions, working memory and ideomotor apraxia in humans ^57^. The Arcuate Fasciculus (AF) in humans is thought to follow a similar trajectory to SFL 2 and have similar functions ^55^. A large metanalysis ^56^ found that the SLF 1 carries primarily spatial information, the SLF 3 encodes non-spatial motor information ^56^, while SLF 2 holds both types of content. VSTM in that study was represented by spatial and non-spatial clusters, and therefore all three tracts could carry VSTM information. In healthy individuals, right predominance of the SLF has been associated with superior performance on visuospatial tasks ^58^. Misbinding errors, as estimated by the mixture model, have been found to be associated with white matter integrity in the SLF (1,2,3), inferior fronto-occipital fasciculus and inferior longitudinal fasciculus in young adults, while in the same study a global reduction of axonal diffusivity in the same tracts was associated with lower memory precision ^59^. We propose that the left SLF 1 and 2, and to a much lesser extent SLF 3 and AF, play a role in the mechanisms underlying the correct representation of an item in VSTM and are also essential to execute the action of selecting the correct item. In healthy controls this function is carried out uniquely by the right SLF 1.

The Superior Thalamic Radiation (STR), or thalamo-parietal bundle, connects anterior and intermediate ventro-lateral (VL) nuclei of the thalamus to the prefrontal cortex, and ventral postero-lateral (VPL) nuclei to the parietal cortex ^60^. These are involved respectively in coordination and planning of movement (VL) and conveying somatosensory input from different body areas (VPL) ^60^. Our data suggests that, similarly to SLF, disruption of the left STR in AD patients is associated with worse ability to hold the representation of an object in VSTM (Identification accuracy) and execute the correct action to select it (Identification time) (**Figure 1-5**, **Table 2-5**).

Lastly, we found that the integrity of the cingulum is also important for VSTM performance in AD patients. In our sample, a disruption of posterior sections of the cingulum was associated with worse Identification accuracy and Target Detection, while the temporal sections showed a correlation with Identification Time (**Figure 2 and 3**, **Table 2, 4 and 5**). The prominent involvement of the Cingulum in AD pathology has been shown by multiple studies ^61, 62, 63^ and also by our own group contrast found in Supplementary materials (**Figure 1S**, **Table 1S and 2S**). However, for the first time we were able to link specific lesions in the posterior sections to maintaining the correct information of the identity of an object in VSTM (Identification accuracy, Target Detection), while temporal sections might be more important to determine the time taken to evaluate and carry out the response (Identification time).

This study has some limitations. The sample size, fairly typical for brain-behavioural studies, is relatively small, and replication in a large dataset would be advisable. Moreover, not all AD patients had CSF/plasma biomarkers. Further studies looking at correlation between these metrics and amyloid and tau positivity would strengthen the biological interpretation of these results in the context of AD pathophysiology. Finally, further studies looking at other measures such as radial, axial and mean diffusivity, or using other models such as neurite orientation dispersion and density imaging (NODDI)^64^, would be advisable to aid microstructural mapping of specific VSTM measures.

## Conclusion

The analysis of white matter integrity reveals that a core area, comprising the left Optic Radiation, Forceps major, Middle Longitudinal Fasciculus is important across different VSTM metrics in patients with AD. In healthy elderly controls a homologous region in the right hemisphere might be important in the processes required for correctly identifying an item. Additionally, specific VSTM functions, such as holding the correct item in memory and selecting it, might require the integrity of the left Superior Longitudinal Fasciculus, Superior Thalamic Radiation, Arcuate Fasciculus and Cingulum in AD.

## Acknowledgments

This research was supported by funding from the Wellcome Trust (206330/Z/17/Z) and National Institute for Health and Care Research (NIHR) Oxford Health Biomedical Research Centre. Y.A.T. was supported by a PhD scholarship by the Friedrich-Ebert-Stiftung. I.M.I was supported by the University of Oxford and the University of Malaya. The funder played no role in study design, data collection, analysis and interpretation of data, or the writing of this manuscript.

## Data availability

De-identified data supporting this study may be shared based on reasonable written requests to the corresponding author. Access to de-identified data will require a Data Access Agreement and IRB clearance, which will be considered by the institutions who provided the data for this research.

