## Supplementary material for "Performance at digital testing in Alzheimer’s Disease is predicted by selective disruption of microstructural integrity"

**Group difference**

We assessed whether our sample showed a main effect of group in line with previous literature, where it has been reported that there is a reduced FA in AD patients compared to healthy elderly controls in several white matter tracts ^12,^ ^65^. AD patients had lower FA in multiple white matter bundles including the Optic radiation (Fornix in JHU atlases), the temporal section of the Cingulum, Forceps major, Forceps minor, Superior and Inferior Longitudinal Fasciculus, Middle Longitudinal Fasciculus, Superior and Inferior Fronto-occipital Fasciculus, Anterior thalamic radiation, Vertical Occipital Fasciculus and Corticospinal Tract (**Figure 1S**, **Table 1S, 2S**). The two biggest clusters, with maximum peaks respectively in the left Optic Radiation and in the right temporal section of the Cingulum bundle, are shown in **Figure 1S** and **Table 1.S** No areas had lower FA in healthy elderly controls compared to AD patients.

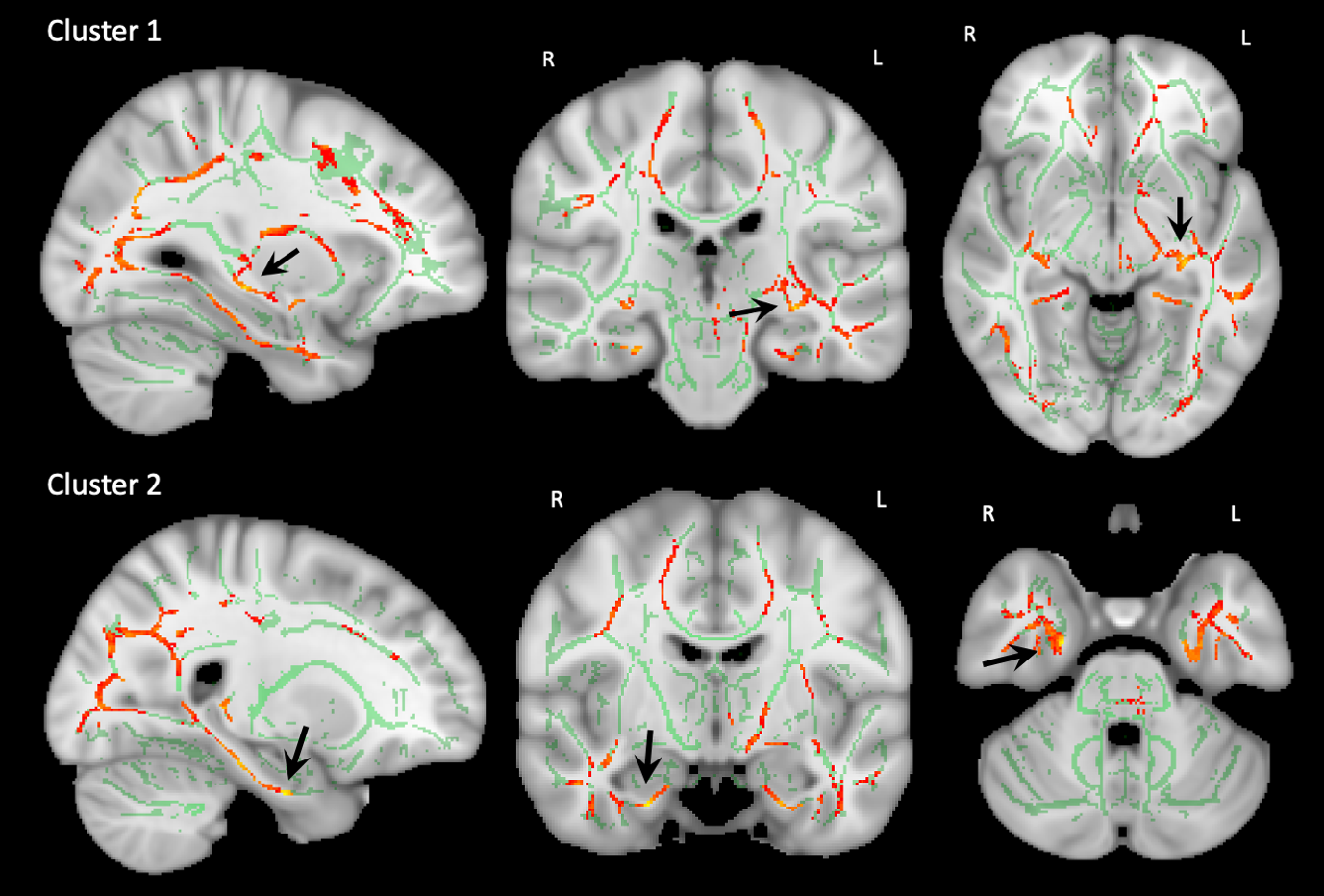

**Figure 1S | Group differences: Clusters 1 and 2**

| **Group** | **White matter tract** | **Cluster Size** | **Coordinates**  **(x,y,z)** | | | **Z-score** |
| --- | --- | --- | --- | --- | --- | --- |
| **1^st^** | Optic Radiation (L) / Fornix (JHU) | 18894 | -30 | -24 | -7 | 5.61 |
| **2^nd^** | Cingulum - temporal section (R) | 11365 | 24 | -9 | -31 | 6.31 |
| **3^rd^** | Forceps minor (L) / Genu of the corpus callosum (JHU) | 1206 | -5 | 23 | -3 | 4.05 |
| **4^th^** | Superior Longitudinal Fasciculus 1 (R) | 864 | 52 | -1 | 19 | 3.9 |
| **5^th^** | Superior Longitudinal Fasciculus 2 (L) | 750 | -31 | 6 | 48 | 4.01 |
| **6^th^** | Fornix (R) | 613 | 26 | -32 | 1 | 5.72 |
| **7^th^** | Forceps minor (R) | 175 | 17 | 43 | -8 | 3.49 |
| **8^th^** | Inferior Fronto-occipital Fasciculus (L) | 34 | -9 | 35 | 47 | 2.42 |
| **9^th^** | Corticospinal tract (L) | 19 | -3 | -33 | -44 | 2.41 |

**Table 1S | Group comparisons: Clusters and diffusivity peaks**

|  | **White matter tract** | **%** |
| --- | --- | --- |
| **Group** | Forceps major | 3.5691 |
|  | Optic Radiation (L) | 3.0312 |
|  | Inferior Fronto-occipital Fasciculus (L) | 2.9021 |
|  | Optic Radiation (R) | 2.2746 |
|  | Superior Longitudinal Fasciculus 1 (L) | 1.9891 |
|  | Cingulum – Temporal section (L) | 1.9024 |
|  | Corticospinal Tract (L) | 1.7636 |
|  | Forceps minor | 1.7201 |
|  | Middle Longitudinal Fasciculus (L) | 1.6679 |
|  | Superior Fronto-occipital Fasciculus 2 (L) | 1.6245 |
|  | Inferior Fronto-occipital Fasciculus (R) | 1.6051 |
|  | Inferior Longitudinal Fasciculus (L) | 1.5985 |
|  | Superior Longitudinal Fasciculus 1 (R) | 1.341 |
|  | Anterior thalamic radiation (L) | 1.3328 |
|  | Middle Longitudinal Fasciculus (R) | 1.2442 |
|  | Cingulum – Temporal section (R) | 1.1725 |
|  | Superior Thalamic Radiation (L) | 1.0652 |
|  | Vertical Occipital Fasciculus (R) | 1.0474 |
|  | Superior Longitudinal Fasciculus 3 (R) | 1.0273 |

**Table 2S | Group comparisons: Tracts probability**
